# Whether Early Steroid dose is associated with lower mortality in COVID-19 critically ill Patients- An exploratory chart review

**DOI:** 10.1101/2020.08.19.20171868

**Authors:** Abhishek Goyal, Saurabh Saigal, Ankur Joshi, Dodda Brahmam, Yogesh Niwariya, Alkesh Khurana, Pooja Singh, Sunaina Tejpal Kama, Jaiprakash Sharma, Sagar Khadanga, G Sai Pavan, Arun Mitra

**Affiliations:** This work was done at AIIMS Bhopal, India

## Abstract

**Introduction:** Steroids have shown its usefulness in critically ill COVID19 patients. However time of starting steroid and dose tailored to severity remains a matter of inquiry due to still emerging evidences and wide-ranging concerns of benefits and harms. We did a retrospective record analysis in an apex teaching hospital ICU setting to explore optimal doses and duration of steroid therapy which can decrease mortality.

**Methodology:** 114 adults with COVID19-ARDS admitted to ICU between 20^th^March-15^th^August2020 were included in chart review. We did preliminary exploratory analysis(rooted in steroid therapy matrix categorized by dose and duration) to understand the effect of several covariates on survival. This was followed by univariate and multivariate Cox proportion hazard regression analysis and model diagnostics.

**Results:** Exploratory analysis and visualization indicated age, optimal steroid, severity (measured in P/F) of disease and infection status as potential covariates for survival. Univariate cox regression analysis showed significant positive association of age>60 years{2.6 (1.5-4.7)} and protective effect of optimum steroid{0.38(0.2-0.72)} on death (hazard) in critically ill patients. Multivariate cox regression analysis after adjusting effect of age showed protective effect of optimum steroid on hazard defined as death {0.46(0.23-0.87),LR=17.04,(p=2e- 04)}.The concordance was 0.70 and model diagnostics fulfilled the assumption criteria for proportional hazard model.

**Conclusion:** Optimal dose steroid as per defined ‘optimum’ (<24 hours and doses tailored to P/F at presentation) criteria can offer protective effect from mortality which persists after adjusting for age. This protective effect was not found to be negatively influenced by the risk of infection.

No funding was taken for this paper.

## Introduction

Since the start of COVID19 pandemic, multiple repurposed drugs like antivirals [Hydroxycholoroquine, Remdesivir], anti-inflammatory (steroids, Tocilizumab or anakinra), anticoagulants and fibrinolytics have been used with variable success.^1,2^ As the pathophysiological pathways around COVID-19 are still advancing in the light of new acquired evidences, one may witness the differential standardized protocols adopted in different settings and gradually evolving protocol in the same setting longitudinally.

The hallmark of COVID-19 associated severe disease is increasing oxygen requirement and increasing C-reactive protein (CRP) levels, which is hypothesized to result from the cytokine storm syndrome (CSS).^3^ This CSS probably acts by two ways: First, it leads to pneumonia and ultimately adult respiratory disease syndrome (ARDS) and second it leads to overreactive coagulation pathway, leading to microvascular thrombosis in the pulmonary vasculature. This dual brunt is the main reason for elevated morbidity and mortality.

At the start of the pandemic, Tocilizumab, an IL6 receptor antagonists had shown benefit in early case series.^4^ But it was not available for most of our patients at our centre because of prohibitive cost ($600-1200 per patient). Pulse dose steroids were used to treat Haemophagocytic lymphohistocytosis (HLH) long before other IL6 antagonists were invented.^5^ COVID19 associated CSS is strikingly similar to HLH in some aspects.^6^ Keeping this scientific pathophysiological plausibility in mind, we decided to give high doses of steroids to patients of COVID19 ARDS. With the context of strong pathophysiological linking of potential steroid therapy amongst COVID19 patients and absence of any standardised treatment protocol at that time, motivated us to initiate steroid therapy amongst COVID19 ARDS patients. As this was a relatively naïve paradigm, standardisation of the dose could not be achieved despite of a qualitative consensus on steroid therapy.

Early administration of glucocorticoid in course of ARDS was associated with better survival in subgroup analysis of patients with SARS/MERS.^7^ Recently published RECOVERY trial has shown positive effect of low dose glucocorticoids (6 mg dexamethasone OD for 10 days) on survival in treating COVID-19 ARDS patients on respiratory support.^8^ But there has been no consensus of usefulness of high dose steroids in literature in COVID19, since pulse steroids is theoretically associated with secondary infections.

We did retrospective chart analysis for patients admitted with COVID19 related ARDS in our ICU, to see whether steroids are associated with reduced mortality and whether it was associated with increased risk of infections.

## Methods

Consecutive adults aged ≥18 years with nasopharyngeal swab PCR-confirmed SARS-CoV-2 infection with ARDS admitted (in ICU) between 20^th^ March,2020 to 15^th^ August 2020 were included in chart review. This review was done in a single tertiary care teaching hospital in central India. The records were reviewed longitudinally from admission to outcome (either till death or discharge) from hospital.

### Definitions

We included only those patients who were given steroids in our ICU during the study period (Figure 1). All our patients in this analysis were being given varying doses of steroids. Time when patient had mMRC grade IV (breathlessness at rest) is routinely noted in all patient’s history. The time difference between onset of severe breathlessness (mMRC grade IV) and first dose of steroid administration (as confirmed by the nursing chart) was counted. Dose and timing of steroid administration was decided by the consultant in-charge posted in ICU.

**Figure1:**
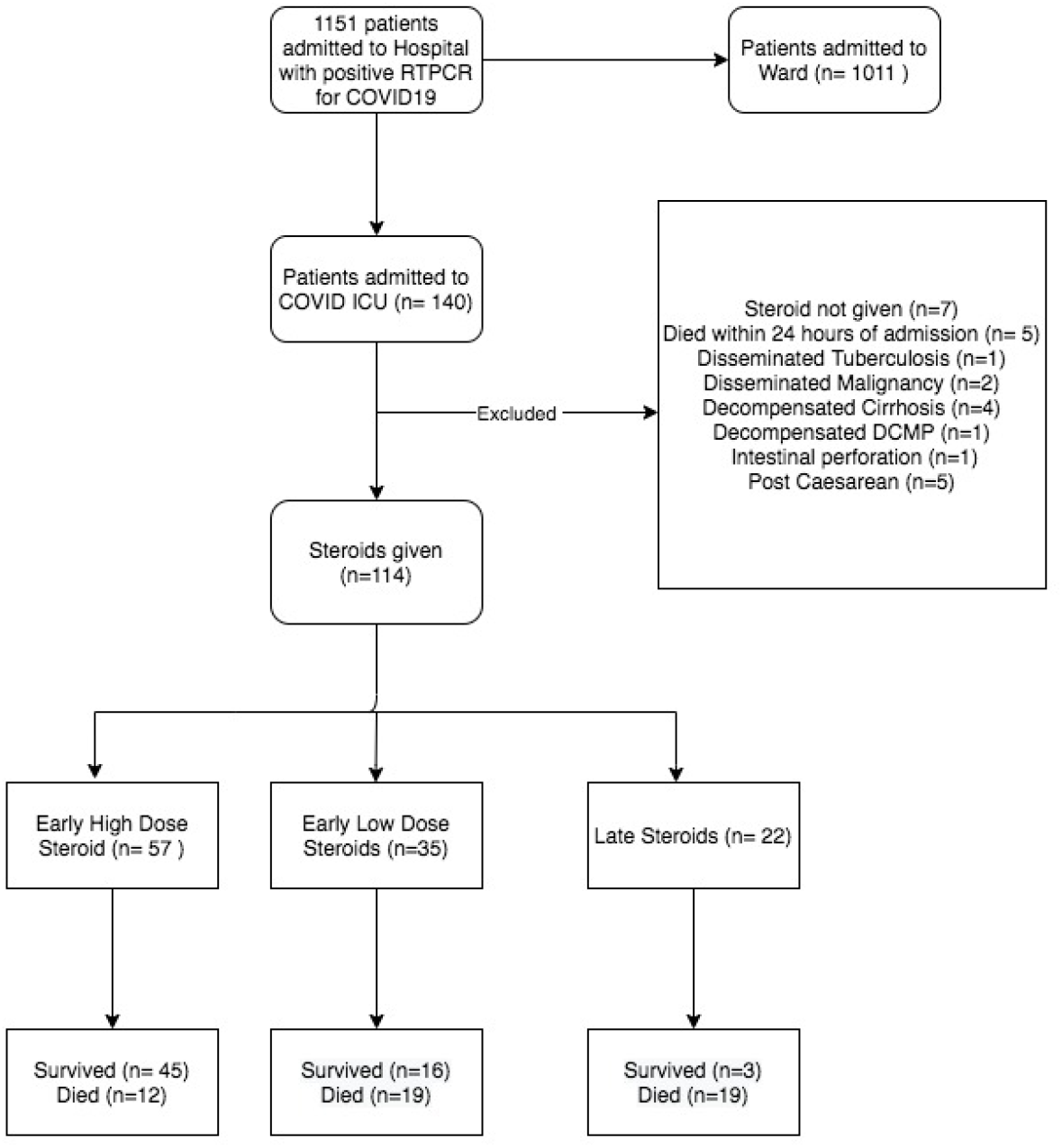
Flowchart of patients admitted in COVID ICU with ARDS

Disease severity was decided on the basis of PaO_2_/FiO_2_ ratio (P/F) on the day of admission to ICU. All patients admitted in our ICU had P/F < 300. For analysis purpose, we categorize these patients into two groups: P/F <150 (critically ill) and P/F in the range of 150-300 (mild to moderate ARDS). Requirement of NIV or invasive mechanical ventilation (IMV) or High Flow Nasal Canula (HFNC) was noted.

As we gained experience in management, we found that high dose steroids (>500 Methylprednisolone) for 2-3 days followed by low dose (30-60 mg MPS) was better in management of patients with high oxygen requirement on admission (P/F <150). For patients with P/F between150 and 300, lower dose steroids (30-125 mg MPS) were sufficient to control cytokine storm. For comparative analysis, we grouped patients into two categories: Optimal and Suboptimal.

We defined Optimal steroids (Table1) as either:

a. If PaO_2_/FiO_2_ (P/F) was less than 150 on Day of admission and patient was given 500 mg of Methylprednisolone (MPS) or more as first day dose of steroid within 24 hours of severe breathlessness (grade IV mMRC). This group is henceforth called as Early Pulse Steroids (EPS).
b. If P/F was more than 150 on day of admission and patient was given 30-125 mg of MPS as first day dose of steroid within 24 hours of severe breathlessness. This group is henceforth called as Early Low Steroids (ELS).

Patients were defined in the suboptimal steroids group (Table 1) if:

I. If first dose of MPS (in any dose) was started after 24 hours of severe breathlessness i.e grade IVmMRC (late steroids). Or
II. If P/F was less than 150 on Day of admission and patient was given less than 500 mg of MPS as first day dose of steroids.

**Table 1:** Definition of Steroids on basis of timing and dose

| Time from grade IV SOB (mMRC) and steroid administration | PaO<sub>2</sub>/FiO<sub>2</sub> on Day 1 | Dose on First day of admission (Methylprednisolone) | Steroid Dose Label | Nested Interpretation |
| --- | --- | --- | --- | --- |
| < 24 hours | >150 | 30 mg to 125 mg | Early Low dose | Optimal Dose |
| < 24 hours | <150 | 30 mg to 125 mg | Early Low dose | Suboptimal dose |
| < 24 hours | <150 | ≥500 mg | Early High Dose | Optimal Dose |
| >24 hours | Any P/F ratio | Any dose | Late steroids | Suboptimal dose |

Process of data extraction- Data was extracted from the case records and treatment files of patients admitted in ICU. Data was retrieved by resident and consultant posted in ICU and was cross checked by another consultant. Data collection included timepoints like symptom onset, onset of severe breathlessness (mMRC grade IV), timing of steroid administration with respect to onset of severe breathlessness, date of hospitalization, date of outcome (discharge or death). Time of grade IV mMRC (breathlessness at rest) is asked from all patients at the time of admission to our ICU as a general practice. Median value of SpO2 was taken for analysis for a 24-hour period. CRP, P/F ratio, FiO2 requirement and absolute lymphocyte counts were recorded and trend was analysed. Death was assessed during hospitalization and patients were followed until discharge from hospital.

All patients in this analysis had hypoxemia and were started on steroids along with empiric antibiotics (for gram negative coverage) to prevent infections. Blood, urine and endotracheal aspirate culture sensitivity were sent whenever patient had fever or TLC increased more than 14000/μl after steroid administration. Bacterial infection was diagnosed if patient had any of the culture reports positive. The retrospective chart analysis was cross checked by two senior consultants.

History of known comorbidities like Hypertension, Diabetes Mellitus (DM), Coronary Artery Disease (CAD), Chronic Kidney Disease(CKD), hypothyroidism was noted from history sheet.

### Statistical Analysis

All statistical analysis was performed using statistical programming language R.^9^ The data was prepared for analysis using standard data cleaning and data wrangling procedures using base R and the ‘tidyverse’ universe of packages. The data cleaning was followed by a comparison of baseline and relevant clinical characteristics among survivors and non survivors through cross-tabulations and bi-variate analysis. Appropriate statistical test (chi square/unpaired t test) were applied. Data visualisation was done using grouped bar charts using functions from the ‘ggplot2’ package.

This exploratory data analysis was tailed by univariate Cox regression analysis for selected variables from exploratory analysis grounded in statistical or clinical significance. Variables with statistically significant regression coefficients were selected for multivariate analysis. A multivariate cox regression analysis was done in order to estimate the effect of optimal steroid category after adjusting the effect of age. The overall robustness of model was assessed by likelihood and wald statistic. The regression coefficients with exponential transformations (point estimates & 95%C.I.) were estimated to obtain hazard ratios. The p value as 0.05 was considered as significant in all analysis.

A forest plot and adjusted survival curve with cumulative hazard function was drawn for visualization of hazard ratios and cumulative survival probabilities.

The model diagnostics were run to check the proportional hazard assumption and presence of influential observations. We did not check for the linearity assumptions as we had only categorical predictor in the model. The model building, visualizations and diagnostics were run with the ‘survival’ and ‘survminer’ package in r -software available in public domain.

Ethics issues-All admitted patients and their relatives are explained about disease condition, therapeutic options and prognosis routinely at our institute. This retrospective chart review study is part of study titled “Predictors of severity in COVID 19 infected patients: Observational Chart review”. Institutional Human Ethics Committee (IHEC) approved the protocol vide LOP Number IHEC-LOP/2020/IM0281, dated 15/07/2020.

Results: During study period, 140 patients were admitted in COVID ICU. Only patients who were RTPCR Positive for COVID19 were included. Twenty six COVID19 patients were excluded because of various reasons.(Figure 1). Total 114 observations (survived=64,died=50) were made for analysis. The relevant baseline characteristics of study population at the time of admission are shown in Table 2.

**Table 2:** Relevant baseline characteristics of study population at the time of admission.

| Characteristic (N=114) | Survived, N = 64<sup>l</sup> | Died, N = 50<sup>l</sup> | $\chi^2$ statistic<sup>#</sup>/ t value* (p value) |
| --- | --- | --- | --- |
| Mean Age ± S.D. in years | 53.16±12.34 | 63.32±12.04 | 4.42 |
| < 60 Years (n=66) | 47 (71.2%) | 19 (29.9%) | **13.0 (p=0.0003)<sup>#</sup>** |
| >60 Years(n=48) | 17 (35.4%) | 31 (64.6%) |  |
| Male (n=89) | 46 (50.5%) | 43 (49.5%) | 2.50 ( p=0.114)<sup>#</sup> |
| Female (n=25) | 18 (64.28%) | 7 (35.7%) |  |
| Diabetes present (n=59) | 32 (54.2%) | 27 (45.7%) | 0.055 (p=0.814)<sup>#</sup> |
| Diabetes absent (n=55) | 32(58.2%) | 23(41.8%) |  |
| Hypertension present (n=56) | 30 (53.6%) | 26 (46.4%) | 0.126 (p=0.72)<sup>#</sup> |
| Hypertension absent (n=58) | 34 (58.6%) | 24 (41.4%) |  |
| Chronic Kidney Disease (n=4) | 2 | 2 | NA |
| Coronary Artery Disease | 6 | 4 | NA |
| ARDS Category on Day 1 |  |  |  |
| 1(n=13) | 10 (76.9%) | 3 (23.1%) | 2.85(p=0.241)<sup>#</sup> |
| 2(n=65) | 36 (55.4%) | 29 (44.6%) |  |
| 3(n=36) | 18 (50%) | 18 (50%) |  |
| P/F ratio on Day 1 | 155.48±54.31 | 131.68±53.26 | **2.35 (p=0.02)<sup>\*</sup>** |
| Absolute Lymphocyte count | 1060±533 | 761±418 | 3.13(p=0.002)<sup>*</sup> |
| Patients on NIV/HFNC (n=87) | 44 | 43 | 3.24 (p=0.07)* |

The next table (Table no.3) shows the distribution of variables pertaining to stay in facility and steroid doses after admission in ICU.

**Table 3.** - Variables of interest after admission in ICU

| Variables | | Outcome | | $\chi^2$ statistic<sup>#</sup>/<br>t value*<br>(p value) |
| --- | --- | --- | --- | --- |
|  |  | Survived | Died |  |
| ICU -Median Length of stay (days) |  | 5(4-9) | 10 (6-14.75) | NA |
| Hospital-Median length of stay (days) |  | 13(10-15) | 10.5 (6.25-15) | NA |
| Infection status- Infected |  | 7 | 16 | **6.48 (p=0.01)<sup>#</sup>** |
| Steroid dose label | Early high (n=57) | 45 | 12 | 29.7 (p<0.0001)<sup>#</sup> |
|  | Early low (n=35) | 16 | 19 |  |
|  | Late (n=22) | 3 | 19 |  |
| Optimal Dose steroid (n=71) |  | 58 | 13 | **47.2 (p<0.0001)<sup>#</sup>** |
| Suboptimal Dose Steroid (n=43) |  | 6 | 37 |  |
| First day Steroid Dose@ |  | 415.78±242.40 | 325.10±265.23 | 1.88 (p=0.06)* |
| Total Steroid Dose @ |  | 1460.78±726.78 | 1459.80±708.98 | 0.007 (p=0.99)* |
| Steroid duration (Days) |  | 7.77± 3.68 | 7.42±4.23 | 0.458(p=0.648)* |
| Tocilizumab (400 mg) (n=8) |  | 5 | 3 | NA |
| Remdesivir (n=15) |  | 7 | 8 | NA |

@ mg Methylprednisolone

This initial exploratory analysis with clinical intuition indicated the role of age category (<60 and >60 years), steroid optimality (defined in Table 1), severity of disease (as measured by P/F) and presence of infections as probable determining factors for the survival. The relationship between severity (P/F status), infection status, age and diabetes status with assumed Optimal steroid dose and outcome is presented in a composite plot in Figure 2. During this study period, invasive mechanical ventilation was used as last resort in our ICU; patients were intubated only when patient could not maintain saturation on NIV or HFNC. We could avoid lot of intubation in patients with P/F<150, in whom early high dose steroids were given. In total, 55 patients were intubated and 9 patients (all 9 were given early high dose) were successfully extubated.

**Figure 2.**
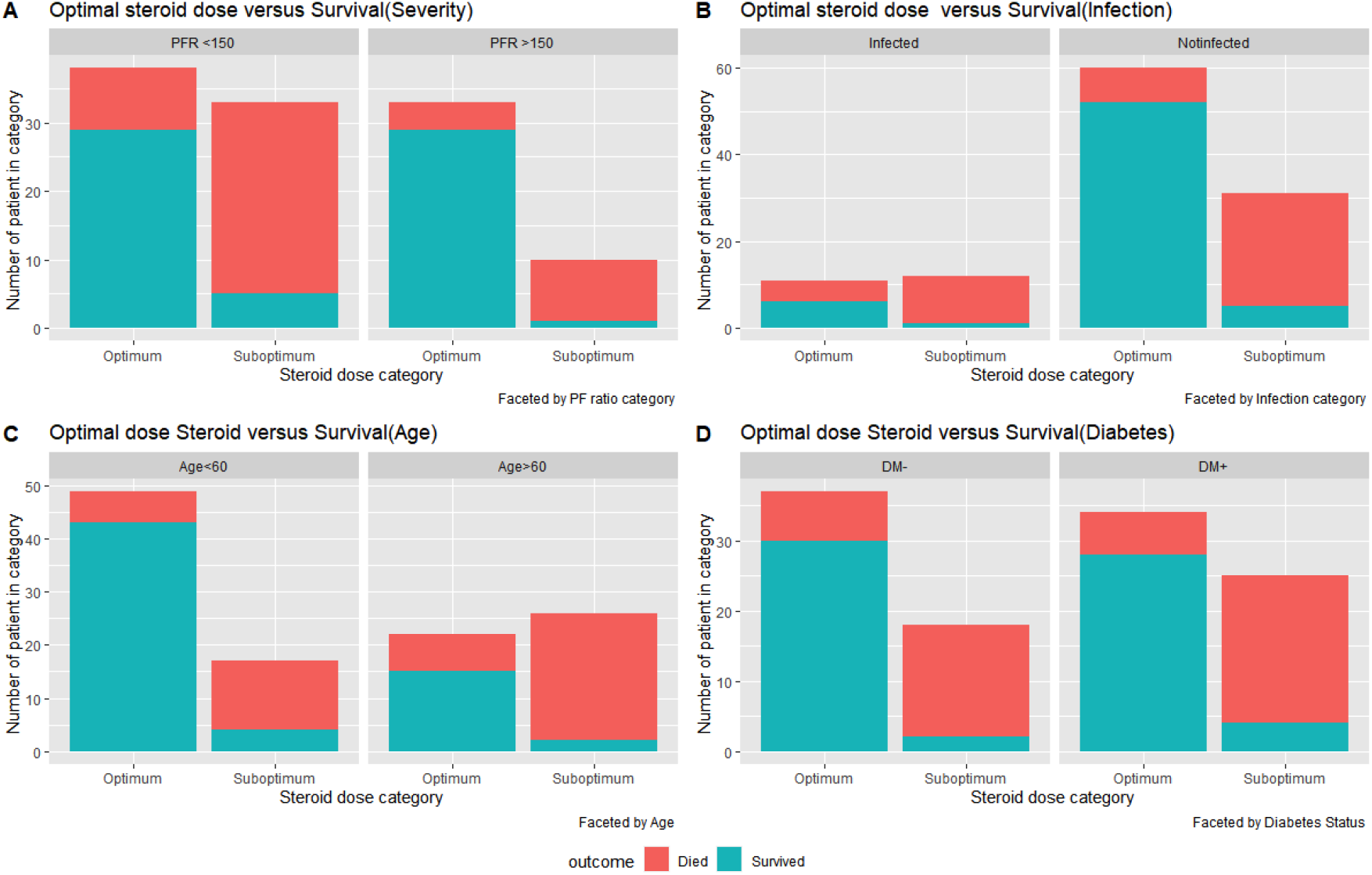
*Composite plot of the distribution of steroid therapy and outcome. The x-axis represents the category of steroid therapy while the y-axis represents the number of study participants in each category. The colour of the bars represent the outcome of the study participants. A: Severity, B: Infection Category, C: Age D: Status of Diabetes*

Figure 2 offers a visual evidence of effect of starting steroid early (<24 hours). Figure-3 is a composite plot of timing and dose of steroid pulse therapy faceted by disease severity as measured in PFR <150 and PFR<100. Figure 3A and 3B may be considered as extension of figure 2. The plot clearly indicate the benefit of early high dose (defined as >500 mg early pulse steroid) in more severe disease(PFR<150). This benefit is more pronounced when P/F cut off of 100 was used as shown in Figure 3B.

**Figure-3:**
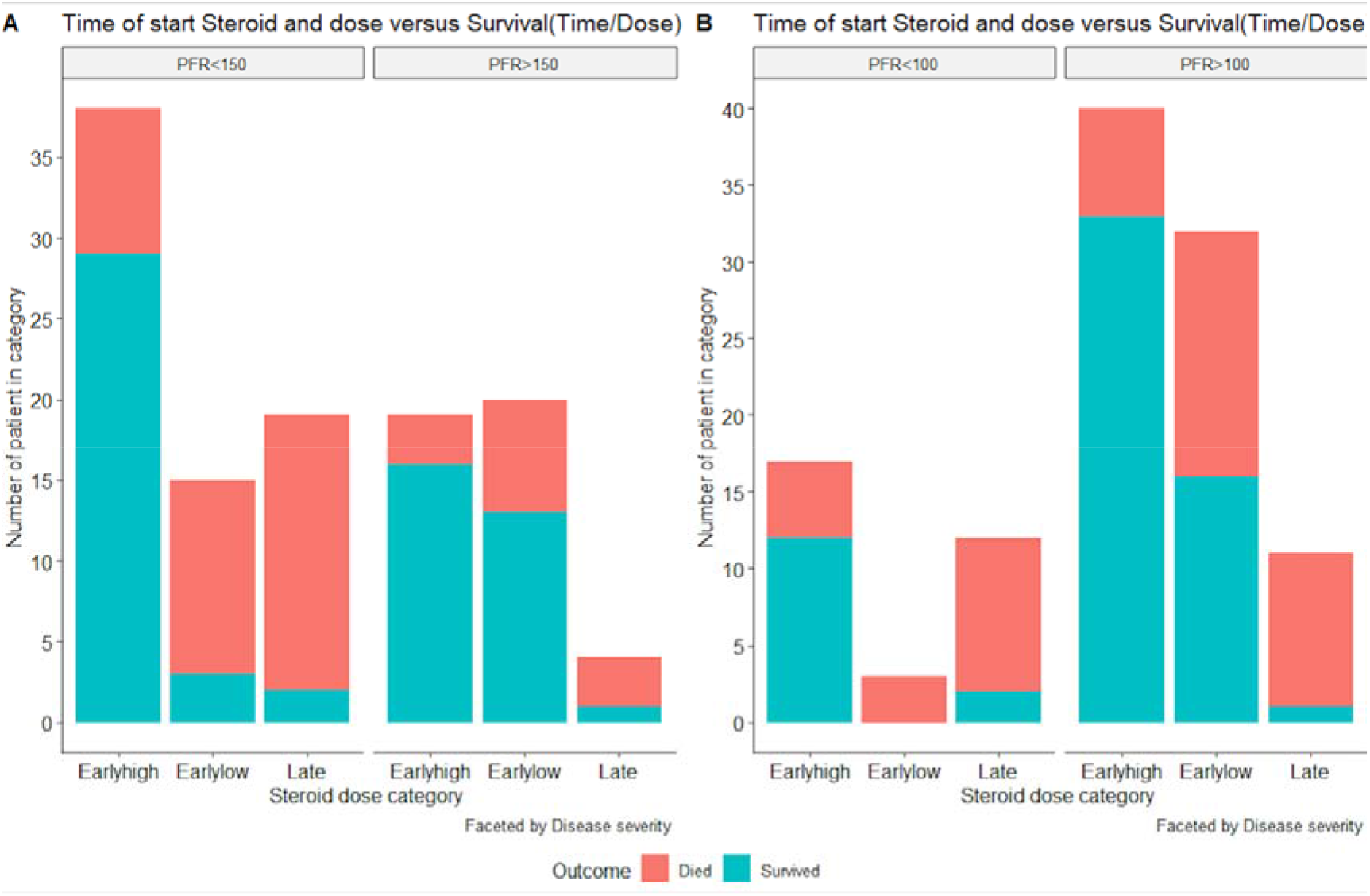
Composite plot of timing and dose of steroid pulse therapy faceted by disease severity as measured in PFR where x-axis represents the category of steroid therapy (which is subdivided into 3 subcategories) while the y-axis represents the number of study participants in each category.

All these variables were further tested through univariate cox regression the result of which is shown in Table-4A.The age>60 years showed a significant positive association with hazard (death) while optimal dose steroid showed a significant negative association with hazard. A multivariate cox regression analysis was done to estimate the effect of optimal steroid category after adjusting the effect of age, the result of which is shown in Table-4B.

**Table 4.**
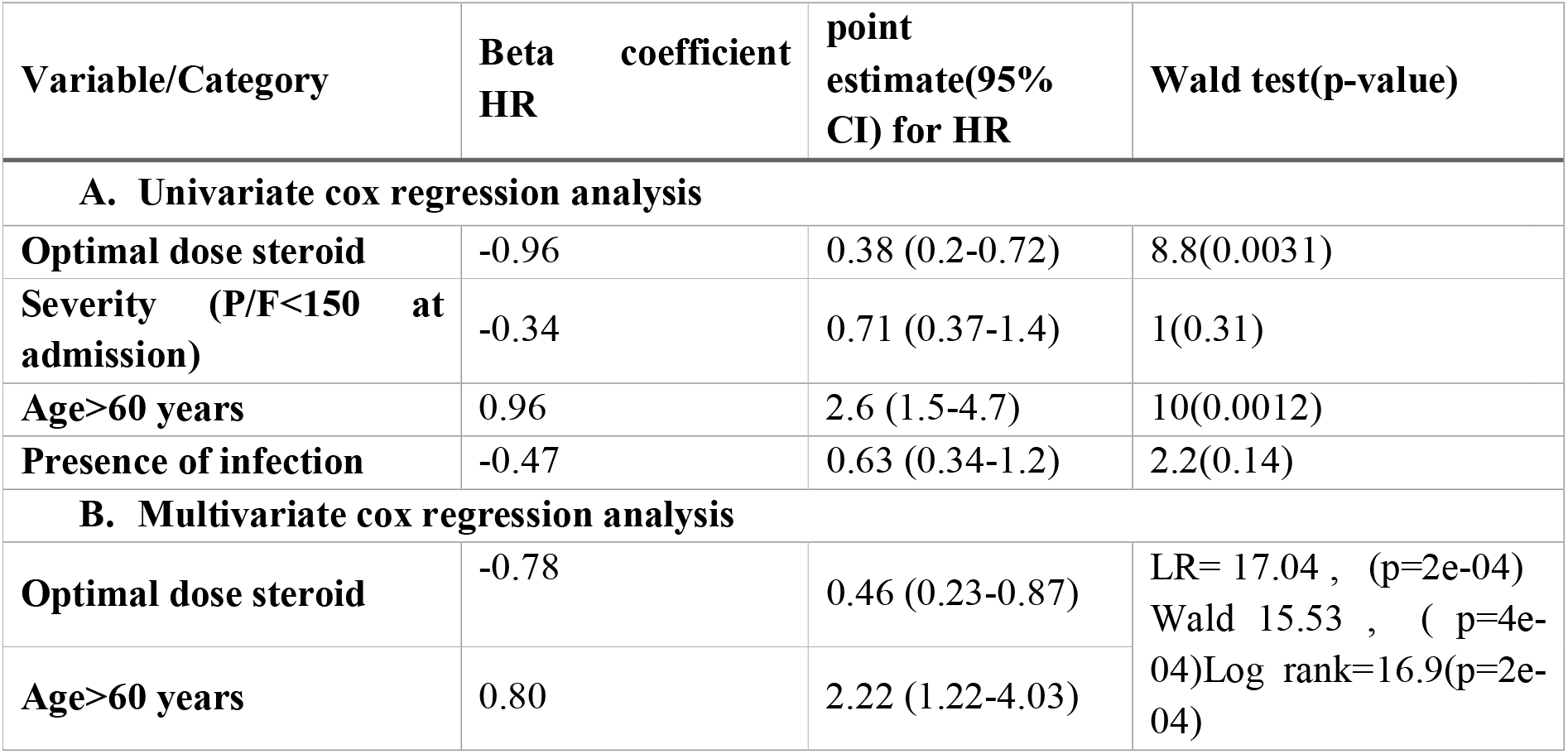
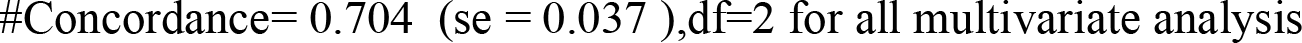
A: Cox regression analysis in univariate (A) and multivariate (B) mode with Hazards Ratio and relevant statistic.

**Fig 4.**
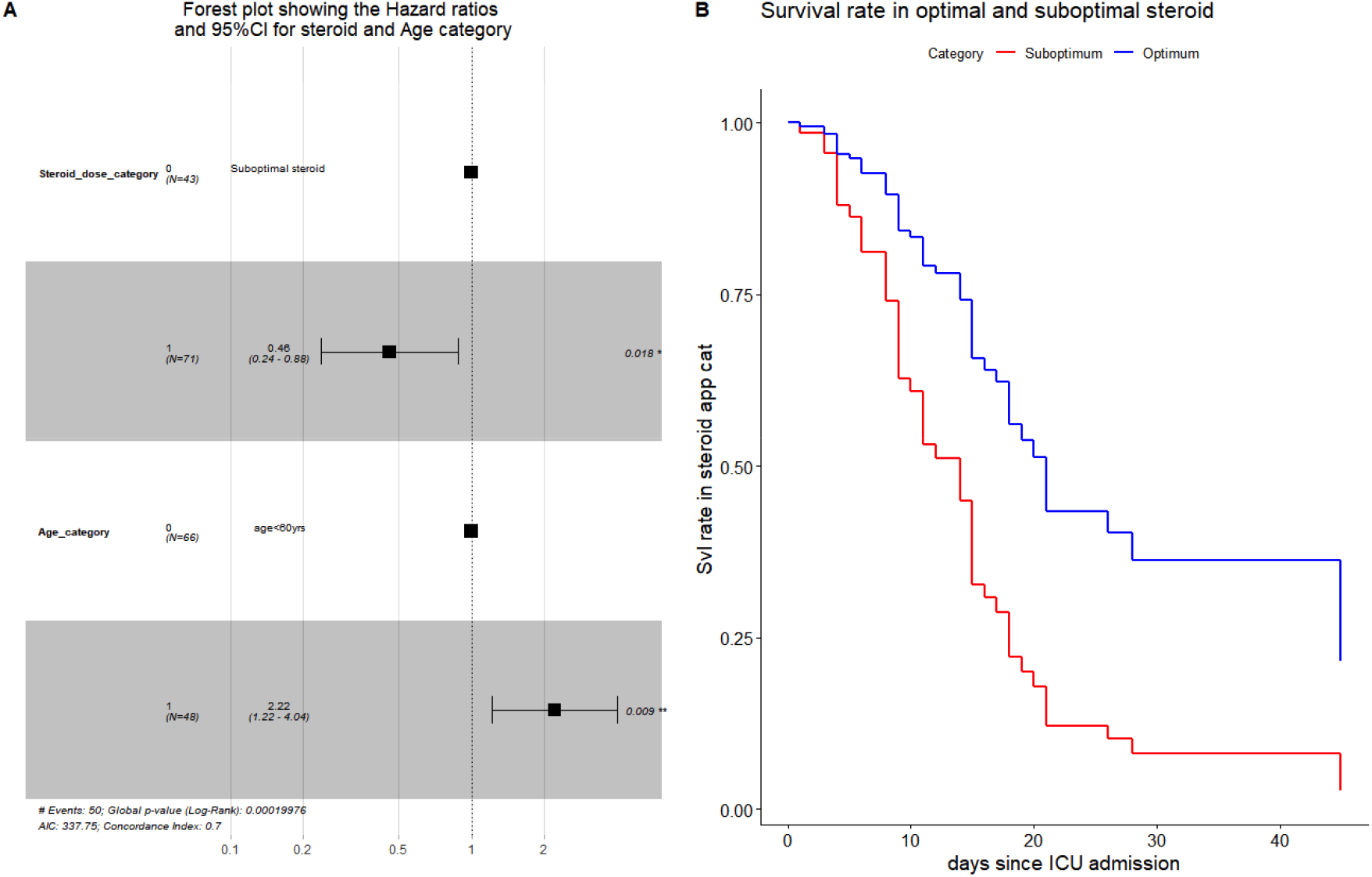
- Forest plot (A) and adjusted survival curve (B) showing the effect of Optimal steroid dose on preventing hazard (death) as estimated by multivariate Cox analysis (0- suboptimal steroid; 1- optimal dose steroid)

The model diagnostics for proportional hazard assumption was supported by an insignificant p values for variables (steroid dose appropriateness = 0.0737,df=1,p=0.79 and age>60 year = 2.1803,df=1,p= 0.14) and for global value of model (2.4881, df= 2 p=0.29). The diagnostic for influential observations was tested by Schoenfeld test for dfbeta residues for variables(age>60years p=0.13,optimal steroid p=0.78)and globally (p=0.78). The test along with plotting the values comparing the magnitudes of the largest df beta against the regression coefficients showed that none of the of the observations was found be excessively influential yet some values for age and Optimal steroid was large compared with the others.

## Discussion

In this retrospective chart analysis study, we found that administration of optimum steroid dose (30 mg to 125 mg in PFR>150 and >500 mg MPS in PFR<150) within 24 hours of start of severe breathlessness was associated with significantly lower mortality in COVID19 ARDS. This phenomenon was not collaterally associated with increase risk of infection..

This protective effect of steroid may be explained with the phasic description of COVID-19 infection timeline-

1. Initial phase of viral replication: Often there are only mild symptoms. Adaptive immunity is stimulated to respond to this replication. In this phase, antivirals (HCQS or Remdesivir) are probably useful. If patient is not on oxygen support, steroids in early phase can be detrimental, as it can increase viral replication.^8^
2. Inflammatory phase: Some patients for currently unknown reasons enter hyperinflammatory phase. This usually starts between 7^th^ to 12^th^ day from initiation of symptoms. It is characterized by varying degree of hypoxemia and breathlessness. Roughly 5-10% of COVID19 patients develop CRS which is characterized by moderate to severe ARDS (PaO_2_/FiO_2_ <200) and elevated CRP/ferritin.^3^ Immunosuppressive (steroids or tocilizumab) have been shown to reduce mortality in inflammatory phase.^4,8,10^
3. Resolution: Most patients with COVID19 do not have heightened immune response to viral replication and they usually do not have any significant symptoms.

Glucocorticoids have been widely used in other viral ARDS like influenza, Severe Acute Respiratory Syndrome (SARS), Middle East respiratory syndrome (MERS), and community-acquired pneumonia with varied success.^7,11^ Low dose steroid (Dexamethasone 6 mg OD equivalent to MPS 30 mg OD) administration during the early inflammatory phase (i.e. Phase 2) has been shown to be beneficial by blunting inflammatory cascade.^8^ In RECOVERY trial benefit was shown in patients on respiratory support (either NIV/HFNC/Oxygen supplementation/ IMV). Authors could not access to the subgroup analysis (if performed) on patients having more severe disease as determined by P/F ratios or oxygen requirement. In a recent metanalysis of 7 prospective trials of steroids in moderate to critical ill patients found survival benefit and has reported pooled odds of 0.70 (0.48-1.01) for mortality.^12^ In this meta analysis, referent group was people who did not receive steroids. But in the trials included in the meta-analysis, cut off for high dose steroids used was 1mg/kg/day for MPS which in our protocol fits to rather low dose category. Whether this difference may be attributed to disparity in severity distribution, settings (ICU versus Non ICU) or other unknown factors is a matter of debate however one can perceive the increasing protective effect of ‘optimum’ steroid dose (>500 mg MPS in PFR<150) as severity increase from PFR<150 to PFR<100. High doses of steroids given early in the phase of hyperinflammation (within 24 hours) can potentially blunt the inflammation cascade if oxygen requirement is high.

In our analysis, steroids were found to be less successful in following conditions:

1. If first dose of steroids was started after 24 hours of severe breathlessness (late steroids) or
2. If low dose steroids were given in patients with initial P/F <150 even if given within 24 hours of severe breathlessness (early low steroids) {Figure 3A and 3B}.

Initially Invasive Mechanical Ventilation was used as last resort in our ICU; patients were intubated only when patient could not maintain saturation on NIV or HFNC. total, 55 patients were intubated and 9 patients (all 9 were given early high dose) were successfully extubated.

Timing of steroids seems to be of importance in patients of COVID19 ARDS. Similar findings were seen in a study from New York, where usage of steroids within 48 hours was associated with better survival.^13^ But in this series, only 10 patients who were given early steroids had sPO_2_<90% on presentation. Also the steroids dose and P/F values were not mentioned in the brief report. In our experience, benefit of high dose steroids seems blunt if given after 2-3 days of grade IV mMRC.

Our assessment of oxygenation shows that improvements in oxygenation starts after 24-36 hours of steroid administration. This was similar to trend seen after tocilizumab infusion seen in a case series, where P/F ratio remains stable for 1-2 days before increasing.^4^ Methylprednisolone is a time tested, readily available, cost effective immunosuppressor unlike new immunosuppressor like tocilizumab, which is cost prohibitive and is available only in selected areas around world. Few RCT comparing steroids and Tocilizumab are underway and results are expected by end of 2020.^10^ We used tocilizumab in only eight patients (3 died and 5 survived) due to prohibitive cost and lack of availability at our centre at the start of pandemic. All eight patients were given high dose steroids plus tocilizumab and six of these developed superimposed fungal infections, gram positive and gram negative bacteraemia. Prescription of tocilizumab along with high dose steroids leads to profound immunosuppression and probably to worse outcome. Remdesivir was given to 15 patients (7 survived and 8 expired) to patients with P/F<150. Three of these 15 patients developed renal failure and all three died.

We experienced rise in total leukocyte count in a significant proportion of patients, but there was no significant difference in rates of infections among pulse and low dose steroid groups. Importantly, presence of infection had insignificant effect on mortality.(Figure 2B) We could detect infection in 20 patients (17 in blood culture, 2 had positive TT asp C/S and one had positive urine C/S). Acinetobacter (n=8), enterococcus (n=5), Enterobacter (n=2),Klebsiella (n=2), Candida (n=2), E Coli(n=1) were the organism found in different cultures. We strongly recommend performing standard of care hygiene precautions in COVID ICU, as these patients are already immunocompromised and giving high dose steroids make them more prone to develop nosocomial infections, which can lead to poor outcome. Protocol followed in our ICU is attached for interested readers. (Supplementary 3)

Another common side effect of high dose steroids is hyperglycaemia, Most of critically ill COVID19 patients are known diabetic, and these patient present with exceptionally raised sugar (sometimes in the range of 400-600 mg/dl) at presentation. Binding of coronavirus to ACE2 receptors found on pancreatic beta cells has been postulated as cause of significant hyperglycaemia.^14^ Markedly raised sugar was present in significant proportion of our patients, but it was easily managed with insulin infusion in all the patients. And survival benefit of early high dose steroids was present even in diabetics.(Figure 2D) So we believe being diabetic should not be impediment for using steroids. Although advanced age was independently associated with poor prognosis, but even in this age group, optimal dose steroids had better results compared to suboptimal dose group. (Figure 2C)

Our study was a retrospective chart analysis from a single centre study with limited sample size. Yet we had maintained a meticulous longitudinal records and have used a robust multivariate methodology and visualization to arrive on outcome. We did not analyse the time taken for RTPCR to become negative. Slower clearance of viral RNA was seen in patients with influenza, SARS and MERS who were treated with glucocorticoids. But significance of persistent positive RTPCR in terms of clinical outcome is unclear in COVID19 ARDS patients who have improved clinically.^15^

In conclusion, optimum dose titrated by disease severity if given early in the course of disease to COVID19 critically ill patients can significantly reduce mortality. A well designed RCT to compare efficacy of low v/s high dose steroids in patients of moderate to severe COVID19 ARDS needs to be conducted to clarify the dose requirement.

## Data Availability

We have submitted data in the supplement.

## Supplementary

Supplementary 1: R Code & complementary analysis

Supplementary 2: Data sheet

Supplementary 3: Existing protocol at our ICU

No funding was taken for this paper.

None of the authors have any conflict of interest.

Concept: AG,SS,AJ

Data Collection: AG,SS, DB, GSP, YN, AK,PS,STK,SK,JS Analysis: AJ,AM

Manuscript preparation: AG,AJ,SS

